# Expected impact of COVID-19 outbreak in a major metropolitan area in Brazil

**DOI:** 10.1101/2020.03.14.20035873

**Authors:** Tarcísio M. Rocha Filho, Fabiana Sherine Ganem dos Santos, Victor Bertollo Gomes, Thiago Augusto Hernandes Rocha, Julio Henrique Rosa Croda, Walter Massa Ramalho, Wildo Navegantes de Araújo

## Abstract

In January 2020 China reported to the World Health Organization an outbreak of pneumonia of undetermined origin in the city of Wuhan, Hubei. In January 30, 2020, the World Health Organization declared the outbreak of COVID-19 as a Public Health Emergency of International Interest (PHEI).

**Objectives:** The aim of this study is to assess the impact of a COVID-19 epidemic in the metropolitan region of São Paulo, Brazil.

**Methods:** We used a generalized SEIR (Susceptibles, Exposed, Infectious, Recovered) model, with additional Hospitalized variables (SEIHR model) and age-stratified structure to analyze the expected time evolution during the onset of the epidemic in the metropolitan area of São Paulo. The model allows to determine the evolution of the number of cases, the number of patients admitted to hospitals and deaths caused by COVID-19. In order to investigate the sensibility of our results with respect to parameter estimation errors we performed Monte Carlo analysis with 100 000 simulations by sampling parameter values from an uniform distribution in the confidence interval.

**Results:** We estimate 1 368 (IQR: 880, 2 407) cases, 301 (22%) in older people (*≥*60 years), 81 (50, 143) hospitalizations, and 14 (9, 26) deaths in the first 30 days, and 38 583 (IQR: 16 698, 113, 163) cases, 8 427 (21.8%) in older people (*≥*60 years), 2181 (914, 6392) hospitalizations, and 397(166, 1205) deaths in the first 60 days.

**Limitations:** We supposed a constant transmission probability *P*_*c*_ among different age-groups, and that every severe and critic case will be hospitalized, as well as that the detection capacity in all the primary healthcare services does not change during the outbreak.

**Conclusion:** Supposing the reported parameters in the literature apply in the city of São Paulo, our study shows that it is expected that the impact of a COVID-19 outbreak will be important, requiring special planning from the authorities. This is the first study for a major metropolitan center in the south hemisphere, and we believe it can provide policy makers with a prognosis of the burden of the pandemic not only in Brazil, but also in other tropical zones, allowing to estimate total cases, hospitalization and deaths, in support to the management of the public health emergence caused by COVID-19.

## Introduction

In January 2020 China reported to the World Health Organization an outbreak of pneumonia of undetermined origin in the city of Wuhan, Hubei. Initially 44 cases were reported, having as common exposure contact the Wuhan seafood market. An increasing number of unrelated secondary cases have since been detected across China, and leading to the dissemination of cases into several countries [1]. The etiologic agent was identified as a new coronavirus, of the betacoronavirus family, which has since be named SARS-CoV-2, and the resulting disease COVID-19 [2]. In January 30, 2020, the World Health Organization declared the outbreak of COVID-19 as a Public Health Emergency of International Interest (ESPII) [3, 4], and in March 11 declared it a Pandemic [5].

Previously to 2019, two highly pathogenic coronavirus had been described in the world. The first named SARS-CoV and described in 2003, was responsible for an epidemic of severe acute respiratory syndrome (SARS), initiated in China and with secondary cases in 26 other countries, accounting for a total of 8,096 cases and 774 deaths [case-fatality rate (CFR): 9.6%] [6, 7]. The second virus, named MERS-CoV, was identified in 2012 is a betacoronavirus responsible for the Middle East Respiratory Syndrome (MERS) [8]. Cases of MERS-CoV are reported sporadically ever since, resulting from zoonotic transmission, with a few outbreaks associated with human to human transmission, resulting in 2449 cases and 845 deaths (CFR: 34,5%), with the majority (84%) reported in Saudi Arabia [9].

SARS-CoV-2 has significant differences relative to MERS-CoV and SARS-CoV. In just over a month of the epidemic, more cases of COVID-19 were confirmed than in the entire history of SARS-CoV and MERS-CoV. Previous outbreaks of SARS-CoV and MERS-CoV have been linked to epidemic amplification phenomena, with few cases were responsible for a disproportionately high number of secondary cases, the so-called super-spreaders, with a significant number of cases resulting from nosocomial transmission. This characteristic allowed outbreaks to occur even in scenarios with an average basic reproduction number *R*_0_ of less than one [4, 10–14]. Unlike SARS-CoV and MERS-CoV, over-dispersion events does not seem to be of a major relevance for the COVID-19 epidemic, suggesting a more homogeneous transmissibility in the population [15, 16].

Homogeneous transmissibility and the potential for transmission from asymptomatic sources [17] brings COVID-19’s behavior closer to other respiratory transmission viruses, such as measles or influenza [13]. Influenza viruses, despite the differences in relation to viruses of the coronavirus family, have similar modes of transmission, and the associated clinical syndromes. Eventually new influenza viruses originating from genetic recombination in animals infect humans, and subsequently transmitted in the population, having been responsible for pandemics in the past, with the occurrence of hundreds of thousands of cases and thousands of deaths worldwide [18, 19].

Brazil has one of the largest public health care systems in the world [20], and understanding how an eventual COVID-19 epidemic in the country could affect this system is central for the preparation of a proper response. In the past, epidemiological models have been used to predict the occurrence of measles cases in order to support decision making in public health emergencies. This paper aims to present tools capable of making projections of the impact of a COVID-19 epidemic in the major metropolitan region of the country. This of particular importance not only for the size of its population, but also for being the main hub of arrival and departure from the country.

We analyze the expected time evolution during the onset of the epidemic in the metropolitan area of São Paulo, with a total estimated population of 21.5 million individuals, the fourth largest in the world. We use a generalized age-stratified SEIR model with the addition of hospitalized population variables (SEIHR model) to predict the occurrence of cases, the expected number of patients admitted to hospitals and deaths caused by COVID-19. Our approach can be adapted straightforwardly to other cities and countries, which is of great relevance in low and middle income countries, with reduced availability of health infrastructure and preparedness to respond to an emergency.

## Materials and methods

It has been shown that age-specific contact rates describes with more accuracy the dynamics of transmission of measles when using an age stratified SEIR model [21], and allows to grasp specifics of social behavior as coded in the contact matrix. Owing to different impacts of the disease across the population according to age, we consider the following age groups: 0–9, 10–39, 40–49, 50–59, 60–69, *≥* 70 years. The population in each group is obtained from the 2010 Brazilian census, corrected by the estimated population in São Paulo in 2019 (see supporting information), except for ages from 0 to 1 year, where the actual population from birth data was used [22, 23]. The variables in the model for each age class are proportions with respect to the total population at time zero: Susceptibles (*S*_*i*_), Hospitalized (*H*_*i*_) due to COVID-19, Exposed (*E*_*i*_) (in the incubation period and not infectious), Infectious (*I*_*i*_) and Recovered (*R*_*i*_) *B* individuals, *i* = 1, …, *M*, (SEIHR model), with *M* the number of age groups. They are such that at time zero we have 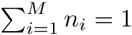 with the fraction of the population in a given age group *n*_*i*_ = *S*_*i*_ + *H*_*i*_ + *E*_*i*_ + *I*_*i*_ + *R*_*i*_ and *M* the number of age groups. The probability of hospitalizations of an infected individual of age-group *i* is estimated as being 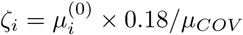 with 0.18 being the proportion of severe and critical cases, *µ*_*COV*_ is the overall estimated letality of the disease and 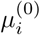 the proportion of fatal cases by the number of cases in age group *i* [24], *which yields ζ*_1_ = 0, *ζ*_2_ = 0.0157, *ζ*_3_ = 0.0313, *ζ*_4_ = 0.102, *ζ*_5_ = 0.282 and *ζ*_6_ = 0.892 for *µ*_*COV*_ = 2.3%. We assume that fatalities only occurs among hospitalized individuals. The different parameter values required are given in current published data and shown withe the corresponding sources in Table 1. The aging rate from age group *i* to age group *i* + 1 is denoted by *ν*_*i*_ and is given by the inverse of the age span of the group (in the corresponding time unit). We put *ν*_0_ = *ν*_*M*_ = 0 in the model equations below.

**Table 1.** Constant parameters in the model.

| Variables | Definition | Value (CI 95%) [Ref] | Distribution |
| --- | --- | --- | --- |
| $\kappa$ | Birth rate | 0.01416 [25] | — |
| $\mu$ | Overall fatality rate from other causes | 0.00608 [25] | — |
| $\psi$ | Average recovery rate from hospital | 1/17.5 days<sup>-1</sup> [26] | — |
| $P_{sc}$ | Proportion of severe and critical cases | 18% [24] | — |
| $\mu_{COV}$ | Fatality rate due to the disease | 0.4% – 2.9% [24] | Uniform |
| $\theta$ | Fatality rate in hospitalized individuals | $\mu_{COV} / P_{sc}$ | — |
| $\sigma^{-1}$ | Inverse of incubation rate | 5.0 (4.2, 6.0) days [27] | Log-Normal |
| $\gamma^{-1}$ | Inverse of recovery rate of non-hospitalized infectious individuals | 1.61 (0.35, 3.23) days<sup>-1</sup> [28] | Not informed.<br>Assumed Log-Norm. |
| $\zeta_i$ | Probability of hospitalizations for age-group $i$ | (see text) | — |
| $R_0$ | Basic reproduction number for COVID-19 | 2.74 (2.47, 3.03)<br>(see supporting information) | Assumed Uniform |
| $\tau_1$ | Median time from illness onset to hospitalization | 3.3 (2.7, 4.0) [27] | Gamma |
| $\tau_2$ | Average time from illness onset to death | 15.0 (12.8, 17.5) [27] | Log-Normal |

Constant parameters in the model with the average value, Confidence Interval (CI) of 95% if pertinent and statistical distribution of values.

We consider that hospitalized individuals are isolated and do not contribute to the force of infection, defined by

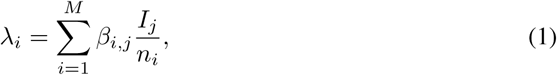

for the *i*-the age group, with *β*_*i,j*_ the transmission matrix, which is estimated as follows. We first consider the contact matrix *C*_*i,j*_ as given by the average number of physical contacts per time unit of and individual of age group *i* with any individual of group *j*. Since the present available information does not allow to determine a probability of contagion for each specific group, we consider the transmission probability per contact *P*_*c*_ being the same for all infected individuals. We thus have that *β*_*i,j*_ = *P*_*c*_*C*_*i,j*_. There are some studies determining the contact matrix for different regions in the world, but none for any Brazilian city. So we considered the study in Ref [29] where the contact matrix was determined from field studies for eight different European countries. Our working hypothesis was that these results can reasonably be transposed for the metropolitan area of São Paulo. Since contact matrices for these different countries do not vary significantly, we take their average for *C*_*i,j*_, and the contact matrix resulting from this procedure is shown as a heat map in Fig 1. The transmission probability is then obtained by adjusting the value of the basic reproduction number from the relation

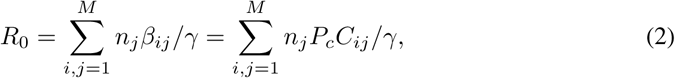

**Fig 1.**
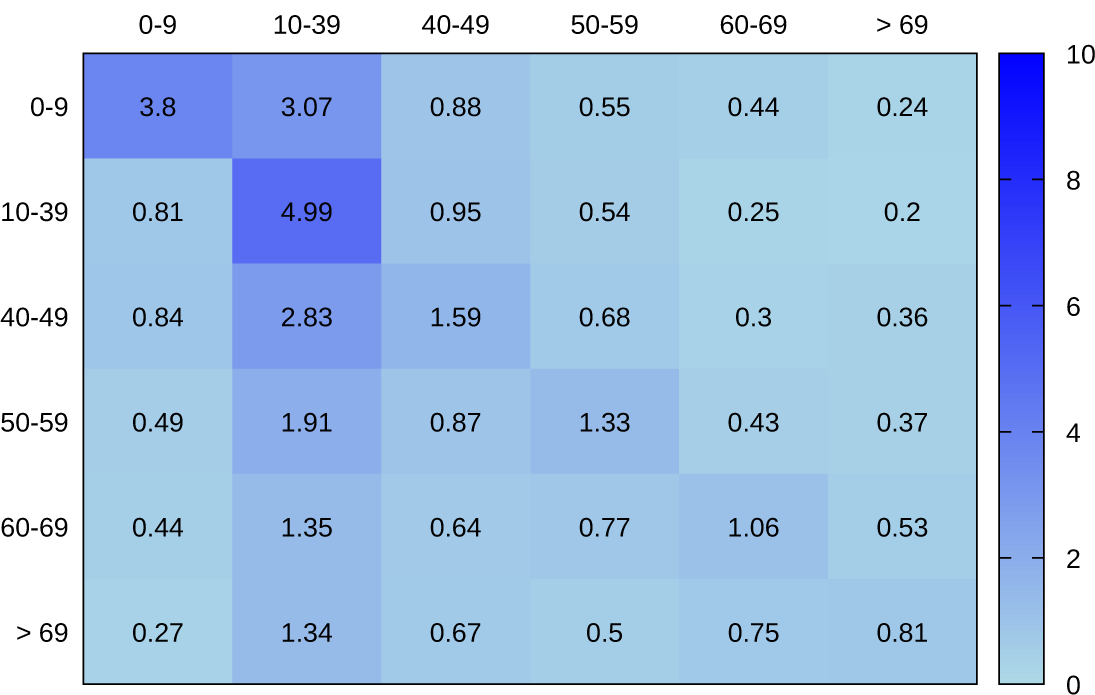
Heat map for the contact matrix *C*_*ij*_ representing the number of physical contacts per day of an individual of age group *i* with any individual of group *j*.

We also suppose that only severe and critical cases are hospitalized.

The model schematic is given in Fig 2 and the corresponding system of differential equation is:

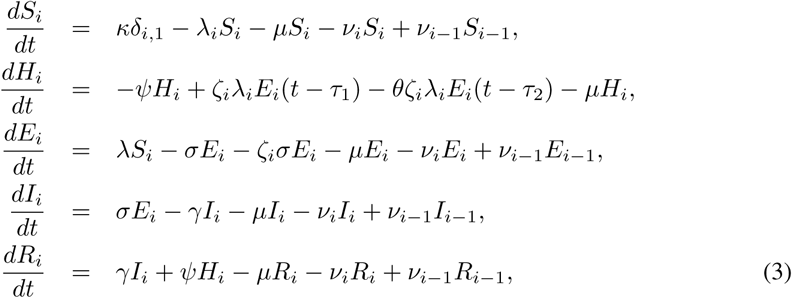

where all variables are taken at time *t* except where explicitly indicated, and *δ*_*ij*_ is the Kronecker delta (1 if *i* = *j* and 0 otherwise). This system is well defined, in the sense that all variables always remain positive and below 1 if there is no population growth. This can be verified straightforwardly simply by noticing that the gradient at the boundaries of the significance region point inward.

**Fig 2.**
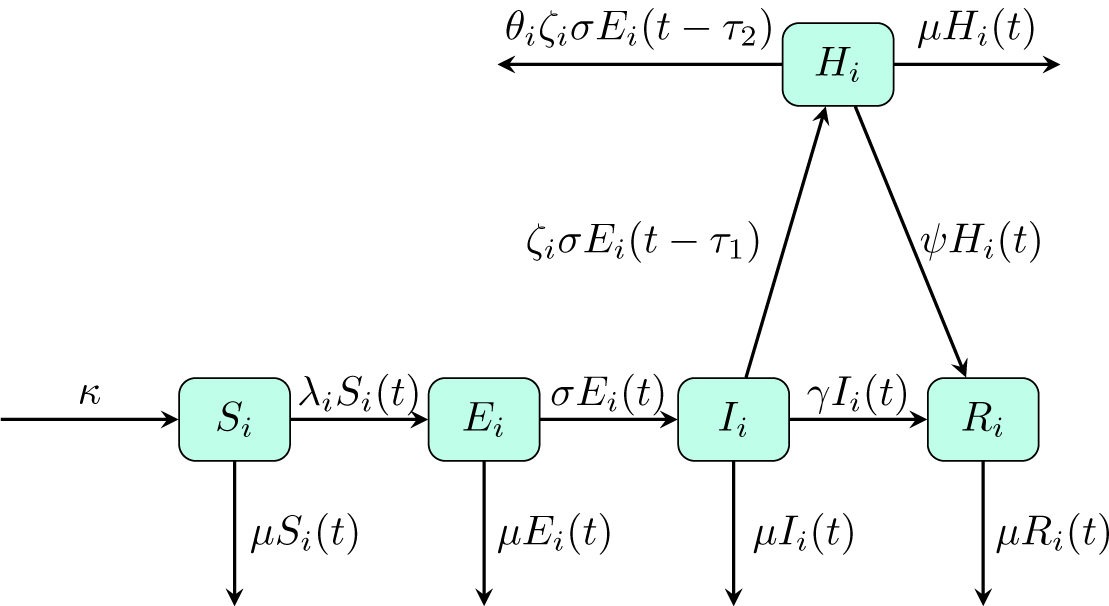
Diagram describing the model equations in Eq (3), constant parameters given in Table 1, and force of infection and transmission rate given in Eqs (2) and (1), respectively.

The solution of Eq (3) was implemented in C, and additional analysis and scripts in the symbolic language system MAPLE and are available on demand.

## Results and discussion

With the onset of an outbreak in a geographically delimited region, a behavioral change is expected in the population in a relatively short time span, as well as government and health officials to intervene with drastic measures in order to reduce contacts, and thence disease propagation. Mathematically, this amounts to judiciously reduce the values of some components of the contact matrix. We consequently restrict ourselves to the first 60 days of the possible outbreak, beyond which results from simulations would not correspond any longer to a realistic setting. Nevertheless, our approach lends itself easily to model specific interventions, as for instance school closure, using the analogous of a school-term forcing in the contact matrix, commonly used to study periodic oscillations in measles [21].

The time evolution for the total number of cases, hospitalized individuals, and total fatalities, for each age group, from the average or median values for the parameters in Table 1, are shown in Figs. 3, 4 and 5, respectively. In order to investigate the sensibility of our results with respect to the estimation errors in different parameter used, we performed a Monte Carlo analysis with 100 000 simulations by sampling parameter values from an uniform distribution, in the interval defined by the corresponding confidence interval. We considered at time zero 10 cases in the age group of 10 to 39 years. In all that follows the number of cases are discounted from this initial value. Results for the medians and inter-quartile intervals for the total number of cases, fatalities and number of hospitalized individual are shown in Table 2, for 30 and 60 days of time evolution. The transmission probability *P*_*c*_ obtained from the Monte Carlos study is well fitted by a log-normal distribution with median of 0.148 and inter-quartile interval of (0.106, 0.247) (see supporting information).

**Table 2.** Median for the total cumulative number of cases averaged over 100, 000 Monte Carlo realizations.

| Age | 30 days |  |  | 60 days |  |  |
| --- | --- | --- | --- | --- | --- | --- |
|  | Cases | Hosp. | Fatal. | Cases | Hosp. | Fatal. |
| 0–9 | 356 (227, 630) | 0 (0, 0) | 0 (0, 0) | 10339 (4493, 30144) | 0 (0, 0) | 0 (0, 0) |
| 10–39 | 299 (196, 519) | 3 (2, 6) | 1 (0, 1) | 8247 (3585, 24115) | 91 (37, 265) | 16 (7, 47) |
| 40–49 | 234 (152, 410) | 5 (3, 10) | 1 (1, 2) | 6587 (2859, 19259) | 141 (58, 410) | 25 (11, 73) |
| 50–59 | 178 (114, 312) | 11 (7, 21) | 2 (1, 3) | 4986 (2156, 14633) | 308 (128, 897) | 55 (24, 163) |
| 60–69 | 151 (96, 267) | 21 (13, 38) | 4 (2, 7) | 4240 (1824, 12522) | 570 (239, 1667) | 103 (44, 312) |
| ≥ 70 | 150 (96, 269) | 39 (25, 69) | 7 (4, 13) | 4187 (1783, 12484) | 1060 (447, 3121) | 196 (80, 605) |
| Total | 1368 (880, 2407) | 81 (50, 143) | 14 (9, 26) | 38583 (16698, 113163) | 2181 (914, 6392) | 397 (166, 1205) |

The number of cases and fatalities are cumulative. The number of hospitalized individuals represent the current number of individuals occupying a place in a hospital setting at this time. The inter-quartile interval is given by the numbers between brackets.

**Fig 3.**
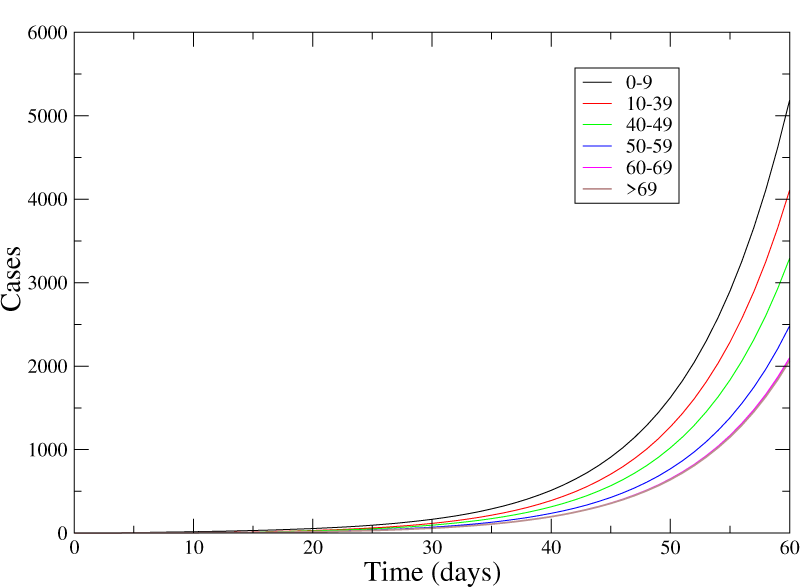
Cumulative number of cases in each age group for the mean or median values for the parameters in Tab. 1, assuming a fatality rate of 2.3%.

**Fig 4.**
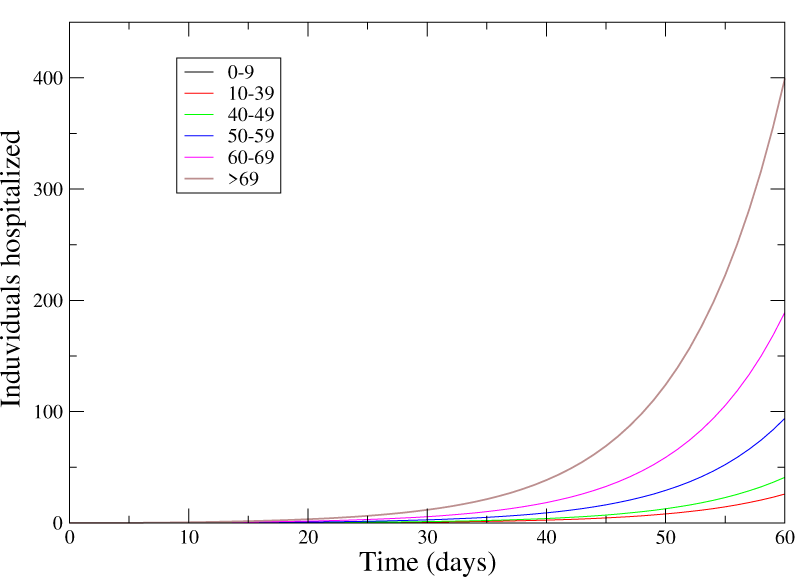
Number of hospitalized individual at a given moment corresponding to Fig 3.

**Fig 5.**
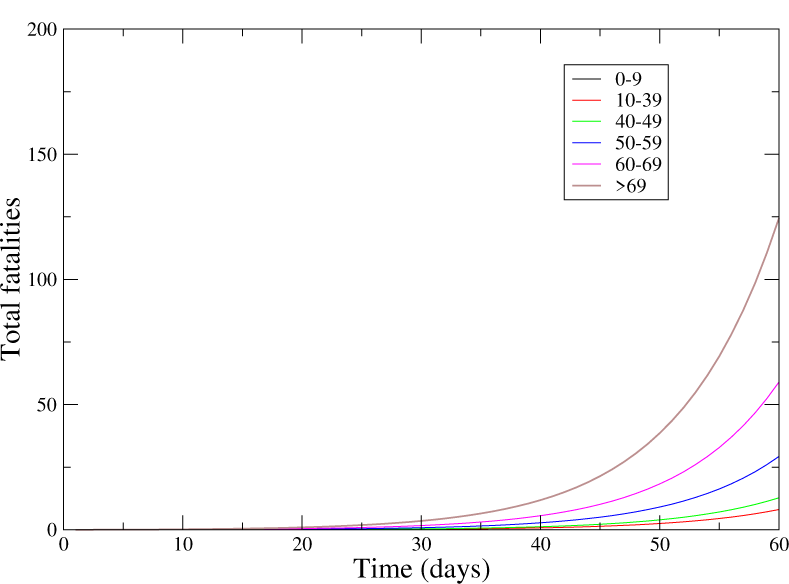
Cumulative number of fatalities corresponding to Fig 3.

This is a first modeling of COVID-19 dynamics using an age-stratified model, similar to approaches for other respiratory diseases epidemics having a historical and clinical relevance in a number of conditions [30, 31]. This type of approach is relevant for the planning of age-dependent intervention policies, e. g. by school closing or restrictions on public gatherings. It is also the first study of a possible epidemic of COVID-19 in a large metropolitan region in the South hemisphere.

Parameters used are described in the literature except for hospitalization probability according to age that was estimated. The contact matrix was adapted from a study of European countries, which are expected to display a similar contact structure as the major city in the southern hemisphere, and comparable in population size to the Wuhan region. Although these two cities have different climates, there is no available empirical evidence to assert its effect on the disease propagation, although clear evidence exists for H1N1 and H3N2 viruses [32]. Further research is in need to clarify this point.

Previous outbreaks of the highly pathogenic coronavirus SARS-CoV and MERS-CoV were related to epidemic amplification with a small number of super-spreader cases causing an elevated number of secondary cases, with a high impact in hospital settings. This explains outbreaks with a basic reproduction number *R*_0_ smaller than one [4, 10–14]. The value of *R*_0_ depends not only on the specifics of the disease, but also on a number of environmental factors, being affected by a change of social behavior in the population and by isolation of infected individuals. This was clearly observed during the recent evolution of the China outbreak, since its onset in December 2019, with a gradual decline of *R*_0_ [33]. This is the main reason why in the present study we restricted the time span of our prognosis to 60 days, as we estimate that after a month of the onset of the outbreak, behavioral change is expected to occur, and public authorities interventions are also expected to occur during the first 60 days from the start of the outbreak.

The present study simulates the impact of COVID-19 on the local health system by a prediction of the number of hospitalized individuals, and consequently provides a tool for policy decision makers to plan the needs of healthcare services in providing people living in an area that is an international hub for the spread of COVID-19, mainly for other countries in South America. The expected number of hospitalized individuals in the first 30 days of the outbreak should be easily absorbed by the existing infrastructure in the metropolitan area of São Paulo, but increases rapidly as the outbreak unfolds, and one expects a rapid saturation of the health settings, which varies according to age. Our approach is therefore a relevant tool to provide authorities responsible for the preparations for a possible outbreak the possibility to know how much time they have at their disposal to prepare complementary health infrastructures. A more systematic and detailed analysis in this directions is the subject of ongoing research.

The determination of the value of *R*_0_ is affected by the supposition that both the time from the start of symptoms, and confirmation, and sub-notification proportions, are roughly constant during the outbreak, resulting in a possible overestimation during the initial stage of the epidemic. The raise in the number of confirmed cases may be in great part be due to a better sensitivity of the health system surveillance and a decrease of the time lag for laboratory confirmation.

## Limitations

In this section we summarize limitations of our work. We assumed that different age-group have the same transmission probability per physical contact, although no information is available to avoid this assumption. We also considered that that every severe or critic case will be in need for hospitalization and supposed, again due to a lack of more detailed data, that the probability of an infected individual from a given age group is proportional to the reported values of the death rate in this same group. Another relevant limitation was to consider that all primary health services will have the same capacity in identifying the severity of clinical conditions, in a region with more than 20 million people living with a GINI index ranging from 0.40 to 0.69 [25], indicating, among other factors, quite different levels of access to tertiary healthcare services. Furthermore, one must consider the possibility that the health infrastructure available can be precociously collapsed. We also assume that, during the 60 days lapse from the epidemic onset, no significant behavioral changes would occur, neither strong interventions from by policy decision makers. More recently the percentage of asymptomatic cases was estimated to be as high as 34.6% [17], while all epidemic parameters previously obtained considered that all cases are symptomatic. On the other hand, the number of asymptomatic cases is expected to have a significant impact on the disease dynamics at later stages, when a substantial proportion was infected. It has nevertheless important consequences in the efficiency of isolation procedures. Although we consider here that hospitalized individuals are no longer able to infect is an oversimplification, and may contribute significantly to the number of reported cases among health professionals. The period that an infected individual transmits the virus, as given in Tab 1, is probably underestimated and was based on the oly available study that explcitly cites it [28].

## Conclusion

Despite having a low case fatality rate, COVID-19 has high transmissibility, with as a consequence a large number of cases when introduced in a naive population, and a large number of hospitalizations and deaths. A COVID-19 epidemic in a major urban area like São Paulo would promote a significant burden in the health care system. Measures to limit the spread of the disease will be necessary in order to slow the epidemic growth and avoid depleting the available hospitals beds and intensive care unit. Mathematical models can contribute in predicting the expected burden of disease. The approach presented here allows a detailed assessment of the impact of the onset of the epidemic in the major metropolitan area in the south hemisphere, despite major limitations in our understanding of the COVID-19 dynamics. For instance, the role of asymptomatic individuals in the progression of the epidemic is still unknown, and no data is available regarding the differences in the transmission rates between different age groups and the impact of the weather in the reproductive number.

## Data Availability

The data will be available on demand

## Acknowledgments

TMRF was partially financed by CNPq (Brazil) under grant no. 305842/2017-0.

## Author contributions

WMR, WNA and JHRC discussed and designed the initial study design. FSGS, VBG and TAHR performed the data gathering and analysis. TMRF wrote and tested all the computer code, run all simulations, and performed the main analysis of corresponding results. Some additional statistical analysis were performed by FSGS and VBG. All authors contributed equally to discussing the final design of the study and participated in the subsequent analysis, discussion and assessment of final results.

## Notes

### Competing Interest Statement

The authors have declared no competing interest.

## References

1. Hui DS, Azhar EI, Madani TA, Ntoumi F, Kock R, Dar O, et al. The continuing 2019-nCoV epidemic threat of novel coronaviruses to global health - the latest 2019 novel coronavirus outbreak in Wuhan, China. Int. J. Infect. Dis. (2020) 91, 264. DOI: 10.1016/j.ijid.2020.01.009.

2. Zhu N, Zhang D, Wang W, Li XW, Yang B, Song JD, et al. A novel coronavirus from patients with pneumonia in China. 2019. N. Engl. J. Med. (2020) 382, 727–733. DOI: 10.1056/NEJMoa2001017.

3. World Health Organization. n. d. Novel Coronavirus (2019-NCoV) Situation Reports. https://www.who.int/emergencies/diseases/novel-coronavirus-2019/situation-reports. World Health Organization. (2004) Summary of Probable SARS Cases with Onset of Illness from 1 November 2002 to 31 July 2003. Emergencies Preparedness, Response. https://www.who.int/csr/sars/country/table2004_04_21/en/. World Health Organization. (2020) Pneumonia of Unknown Cause – China. Disease Outbreak News 2020. https://www.who.int/csr/don/05-january-2020-pneumonia-of-unkown-cause-china/en/.

4. World Health Organization. Consensus Document on the Epidemiology of Severe Acute Respiratory Syndrome (SARS). WHO/CDS/CSR/GAR/2003. 11, 1–47. https://www.who.int/csr/sars/en/WHOconsensus.pdf

5. WHO Director-General’s opening remarks at the media briefing on COVID-19 - 11 March 2020. https://www.who.int/dg/speeches/detail/who-director-general-s-opening-remarks-at-the-media-briefing-on-covid-19—11-march-2020.

6. Drosten C, Günther S, Preise Wr, van der Werf S, Brodt HR, Becker S, Rabenau H et al. Identification of a Novel Coronavirus in Patients with Severe Acute Respiratory Syndrome. New England Journal of Medicine (2003) 348(20): 1967–1976. https://doi.org/10.1056/NEJMoa030747.

7. Zhong NS, Zheng BJ, Li YM, Poon LLM, Xie ZH, Chan KH, Li PH, et al. Epidemiology and Cause of Severe Acute Respiratory Syndrome (SARS) in Guangdong, People’s Republic of China, in February, 2003. The Lancet (2003) 362 (9393): 1353–1358. https://doi.org/10.1016/S0140-6736(03)14630-2.

8. Zaki AM, van Boheemen S, Bestebroer TM, Osterhaus ADME, Fouchier RAM. Isolation of a Novel Coronavirus from a Man with Pneumonia in Saudi Arabia. New England Journal of Medicine (2012) 367 (19): 1814–1820. https://doi.org/10.1056/NEJMoa1211721.

9. World Health Organization. WHO MERS Global Summary and Assessment of Risk. WHO/MERS/RA/August18 (2018). https://www.who.int/csr/disease/coronavirus_infections/risk-assessment-august-2018.pdf.

10. Varia M, Wilson S, Sarwal S, McGeer A, Gournis E, Galanis E, Henry B. Investigation of a Nosocomial Outbreak of Severe Acute Respiratory Syndrome (SARS) in Toronto, Canada. Cmaj (2003) 169 (4): 285–292.

11. Oboho IK, Tomczyk SM, Al-Asmari AM, Banjar AA, Al-Mugti H, Aloraini MS, Alkhaldi KZ et al. 2014 MERS-CoV Outbreak in Jeddah – A Link to Health Care Facilities. New England Journal of Medicine (2015) 372(9): 846–854. DOI: 10.1056/NEJMoa1408636.

12. Jeong D, Lee CH, Choi Y, Kim J. The Daily Computed Weighted Averaging Basic 0,k,Ω Reproduction Number Rn for MERS-CoV in South Korea. Physica A (2016) 451: 190–197. DOI: 10.1016/j.physa.2016.01.072.

13. Chowell G, Abdirizak F, Lee S, Lee J, Jung E, Nishiura H, Viboud C. Transmission Characteristics of MERS and SARS in the Healthcare Setting: A Comparative Study. BMC Medicine (2015) 13:210: 1–12. DOI: 10.1186/s12916-015-0450-0

14. Cowling BJ, Park M, Fang VJ, Wu P, Leung GM, Wu JT. Preliminary Epidemiologic Assessment of MERS-CoV Outbreak in South Korea, May–June 2015. Euro Surveill. (2015) 20(25): 21163. DOI: 10.2807/1560-7917.es2015.20.25.21163

15. Novel Coronavirus Pneumonia Emergency Response Epidemiology Team 2020

16. Riou J, Althaus CL Pattern of Early Human-to-Human Transmission of Wuhan 2019 Novel Coronavirus (2019-NCoV), December 2019 to January 2020. Eurosurveillance (2020) 25(4): 2000058. DOI: 10.2807/1560-7917.ES.2020.25.4.2000058

17. Mizumoto K, Kagaya K, Zarebski A, Chowell G. Estimating the Asymptomatic Ratio of 2019 Novel Coronavirus onboard the Princess Cruises Ship, 2020. medRxiv preprint DOI: 10.1101/2020.02.20.20025866.

18. Kilbourne ED. Influenza Pandemics of the 20th Century. Emerging Infectious Diseases (2006) 12(1): 914. DOI: 10.3201/eid1201.051254.

19. Potter CW. A History of Influenza. Journal of Applied Microbiology (2001) 91(4): 572–79. DOI: 10.1046/j.1365-2672.2001.01492.x.

20. Victora CG, Barreto ML, Leal MC, Monteiro CA, Schmidt MI, Paim J, Bastos FI, Almeida C, Bahia L, Travassos C, Reichenheim M, Barros FC, the Lancet Brazil Series Working Group. Health conditions and health-policy innovations in Brazil: the way forward The Lancet (2011) 377:2042–2053. DOI: 10.1016/S0140-6736(11)60055-X

21. Magpantay FMG, King AA, Rohani P. Age-structure and transient dynamics in epidemiological systems. Journal of the Royal Society Interface (2019) 16: 20190151. DOI: 10.1098/rsif.2019.0151.

22. Instituto Brasileiro de Geografia e Estatística (IBGE). https://cidades.ibge.gov.br/brasil/sp/sao-paulo/panorama

23. http://dive.sc.gov.br/index.php/sistemas-de-informacao/sistemas/pagina-sinasc.

24. The Novel Coronavirus Pneumonia Emergency Response Epidemiology Team. The Epidemiological Characteristics of an Outbreak of 2019 Novel Coronavirus Diseases (COVID-19) — China, 2020. CCDC Weekly (2020) 2(x): 1.

25. Instituto Brasileiro de Geografia e Estatística (IBGE). https://brasilemsintese.ibge.gov.br/populacao/

26. Wang C, Hornby PW, Hayden FG, Gao GF. A novel coronavirus outbreak of global health concern. Lancet (2020) 395, 470. DOI: 10.1016/S0140-6736(20)30185-9.

27. Linton NM, Kobayashi T, Yang Y, Hayashi K, Akhmetzhanov AR, Jung S, Yuan B, Kinoshita R, Nishiura H, Incubation Period and Other Epidemiological Characteristics of 2019 Novel Coronavirus Infections with Right Truncation: A Statistical Analysis of Publicly Available Case Data. Journal of Clinical Medicine (2020) 9: 538.

28. Read JM, Bridgen JRE, Cummings DAT, Ho A, Jewell CP. Novel coronavirus 2019-nCoV: early estimation of epidemiological parameters and epidemic predictions imedRxiv 2020.01.23.20018549; doi: https://doi.org/10.1101/2020.01.23.20018549

29. Mossong J, Hens N, Jit M, Beutels P, Auranen K, Mikolajczyk R, Massari M, Salmaso S, Tomba GS, Wallinga J, Heijne J, Sadkowska-Todys M, Rosinska M, Edmunds WJ. Social Contacts and Mixing Patterns Relevant to the Spread of Infectious Diseases. PLOS Medicine (2008) 5: 74.

30. Hethcote HW. The Mathematics of Infectious Diseases. SIAM Review (2000) 42(4): 599–653.

31. Keeling MJ, Rohani P. Modeling Infectious Diseases in Humans and Animals. Princeton University Press (Princeton, 2008).

32. Steel J, Palese P, Lowen AC, Transmission of a 2009 Pandemic Influenza Virus Shows a Sensitivity to Temperature and Humidity Similar to That of an H3N2 Seasonal Strain. Journal of Virology, 85(3): 1400–1402. DOI: 10.1128/JVI.02186-10.

33. Liu T, Hu J, Xiao J, He G, Kang M, Rong Z, et al. Time-varying transmission dynamics of Novel Coronavirus Pneumonia in China. bioRxiv preprint DOI: 10.1101/2020.01.25.919787.

